# UV light influences covid-19 activity through big data: trade offs between northern subtropical, tropical, and southern subtropical countries

**DOI:** 10.1101/2020.04.30.20086983

**Authors:** Novanto Yudistira, Sutiman B. Sumitro, Alberth Nahas, Nelly Florida Riama

## Abstract

UV (ultraviolet) light is an important factor should be considered to predict coronavirus epidemic growth pace. UV is different from weather temperature since UV is electromagnetic wavelength from 10 nm to 400 nm in size, shorter than of visible lights. For some people, UV light can lead to cancer from unprotected sun exposure, however, for tropical people, which have been used to live in such condition, have resisted from negative effect high UV index. Moreover, UV has the capability to inactivate virus. This conclusion has been discussed deeply with biological experts. Although UV light has the ability to inactivate viruses, it may be meaningless in areas with high air pollution where UV light turns into heat. The data visualization code is available here https://github.com/cbasemaster/uvcorona

## 1 Introduction

Coronavirus disease 2019 (Covid-19) was first detected in Hubei province of China by Wuhan Municipal Health Commission and early information about outbreak has been sent to World Health Organization (WHO) [1][2]. Disease capability to spread among communities are increased rapidly as the number of people that exposed to covid-19 is increasing. This regards evidence of human-to-human transmission indicating that covid-19 is highly contagious. Moreover, covid-19 can also actively live airborne and in surface [3]. These kind of transmissions can form pandemics. Pandemics can cause severe mobility and mortality over wide geographic area [4]. However, the efforts to halt covid-19 spread reduce carbon emission[5] of which advantageous for environment. To withstand spread of covid-19, one natural instrument that rarely discussed is ultraviolet (UV). There are many UV papers prove that it has capability to inactivate virus [6][7]. An example about virus inactivation by UV is viral inactivation using UV-C irradiation [8]. Even though UV has the ability to inactivate the virus but if the pollution is high it will be meaningless [9]. Note that smoke particulate is able to weaken the UV light ability to exist in the air [10]. Moreover, The vaccine development is not effective and taking a long time to be found [11]. Therefore, urgent, massive and natural immune are somewhat more desirable. Previously, some technologies have been developed by making use of UV light [12][13][14]. By aforementioned evidences, we investigate how UV is related to geographical locations and the spread of covid-19 in the world starting from Indonesia. Our assumption is that there is advantage of country like Indonesia where UV index is very high to withstand the spread of covid-19. As information, data sets of world confirmed covid-19, UV index time series, pollution time series, and UV index of Indonesia are gathered from www.github.com/datasets/covid-19/tree/master/data, www.temis.nl/uvradiation/UVarchive [15], www.aqicn.org, and BMKG (Meteorology, Climatology, and Geophysical Agency) www.bmkg.org, respectively during 2020-01-22 to 2020-03-28.

### 1.1. How UV index is determined

The UV index is derived from the measured solar radiation in the UV spectra that arrives on the surface. It is calculated by considering the proportional contribution of both UV-A and UV-B, two of the three wavelength-based types of UV radiation. UV-A is characterized as the UV radiation of which the wavelength ranges from 280-315 nm, while the wavelength of UV-B is between 315 nm and 400 nm.

The determination of UV index stems from the importance of its health impacts to humans. Therefore, this index was developed to assess how the exposure of excessive UV radiation may be detrimental particularly to the exposed body. Both UV-A and UV-B have this damaging capability if the exposure is not being taken cautiously. However, there are some differences when it comes to the health effects by each type of UV radiation.

When determining the UV index, the contribution of UV-A and UVB is translated by implementing the weighting factors for individual UV types. These factors were introduced by [16] to account for the amount of received UV radiation on surface and the attributed energy of each UV type. This level of energy is indicative of how damaging the exposure might be experienced by a person. In general, UV radiation at longer wavelengths is received more abundantly than that at shorter wavelengths. In other words, more UV-A arrives on the surface than UV-B. However, the energy carried by UV-A is less than that by UV-B because of the inverse relationship between the energy amount and the radiation wavelength.

The weighting factors for determining the UV index accommodate both the amount of received UV radiation and carried energy. A study by [16] has provided a list of weighting factors for each nm wavelength of UV-A and UVB, with decreasing values assigned from lower to higher wavelengths (*i.e*., the weighting factors for UV-B are higher than those of UV-A). These factors are utilized to account for how much UV energy received on the surface that might affect the human health.

In the ideal situation, the application of the weighting factors to UV-A and UV-B radiation is performed for each nm of wavelengths. However, data from UV radiation is usually reported as a total of integrated UV radiation from a range of wavelengths. As a result, applying the weighting factors cannot be done to specifically determine the contribution of UV radiation from every wavelength. With this compromised situation, an alternative approach has been undertaken to select a weighting factor for a particular wavelength that represents each UV-A and UV-B wavelength range. Since the chosen wavelength can be arbitrary, some reasonable assumptions were provided to serve as justification of calculation. For UV-A, the weighting factor used was selected for the wavelength of 325 nm (UV-A weighting factor = 0.0029). Although UV radiation from this wavelength penetrates to the surface less than that from longer wavelengths, it has a higher weighting factor that might be considered as a surrogate for other wavelengths.

Meanwhile for UV-B, the selected weighting factor was for 305 nm (UV-B weighting factor = 0.22). This wavelength is located near the end of the UVB spectrum, indicating a lower energy amount than the other shorter UV-B wavelengths. However, much of the shorter wavelengths are already absorbed by gases in the atmosphere before arriving at the surface. Therefore, it can be assumed that the prevalent UV-B radiation received on the surface is from the longer wavelengths. In addition, the weighting factor for 305 nm is located in the middle area of the UV-B longer wavelength spectrum, which is appropriate to represent this range in calculation.

After applying the weighting factors to both UV-A and UV-B, the combined energy is then used to determine the UV index. The UV index is mathematically expressed by the following Eq. (1):

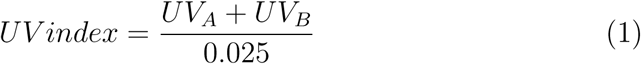

where both UV-A and UV-B are expressed in W m^−2^. The denominator 0.025 W m^−2^ is the standardized increment value that corresponds to how much UV radiation can potentially be damaging to life tissues. In other words, an increase of one UV index is equivalent to 25 mW m^−2^ of exposed UV radiation.

### 1.2. Information of UV index in Indonesia

UV radiation is one of the solar radiation components pertinent in determining the atmospheric dynamics. Therefore, this parameter has been an integral parameter to be measured on the surface. However, there is still a limited number of ground-based measurements of UV radiation in Indonesia. Right now, the Indonesian Agency for Meteorology, Climatology, and Geophysics (BMKG) operates a UV radiometer to measure UV radiation in three climatological stations. These three locations, shown in SM2a, report the incidence UV radiation received on the surface per hour. These values are useful to determine the daytime UV radiation profile in each location.

The limitation of ground-based data for UV radiation poses some challenges in determining UV index for a country like Indonesia. However, this limitation can be addressed by utilizing model-based UV radiation information. Such information, for example, can be obtained from the European Centre for Medium-Range Weather Forecast (ECMWF) UV radiation products. ECMWF releases two near real-time global UV radiation products, which are UV biologically effective dose (uvbed) and UV biologically effective dose for clear sky (uvbed). The horizontal resolution of these products is 0.4by 0.4, which is roughly equivalent to 40 km by 40 km. Both products are adjusted UV radiation received on the surface that take into account atmospheric and surface conditions, such as surface albedo, clouds, aerosol loads, and surface ozone. Their values are analogue to the total weighted UV radiation described earlier. As such, these products can also be converted into a UV index by dividing the values by 0.025 W m^−2^.

The spatial variation of UV index calculated from the ECMWF’s uvbed product for April 15, 2020, is depicted in fig. 1. In this figure, three selected hours (*i.e*., 0 UTC, 5 UTC, and 9 UTC, where Jakarta Time is seven hours ahead of UTC) are shown to observe the changing of UV index from morning to afternoon. At 0 UTC or 7 AM Jakarta Time, the central and eastern parts of Indonesia have observed UV index 1 to 3, while most of the western part was yet to be affected by UV radiation. As the day progressed and the sun was overhead, most of the Indonesian region was under UV index 7 and above by 5 UTC or noon Jakarta time. In the afternoon, as the sun was observed at a slanted angle, the UV index was either decreased or zero. These changes of UV index are typical for daily observation. There are some seasonal variations in terms of magnitude that is related to the geometry between the sun and the earth. In addition, surface conditions and atmospheric dynamics contribute to determining the UV index.

**Figure 1:**
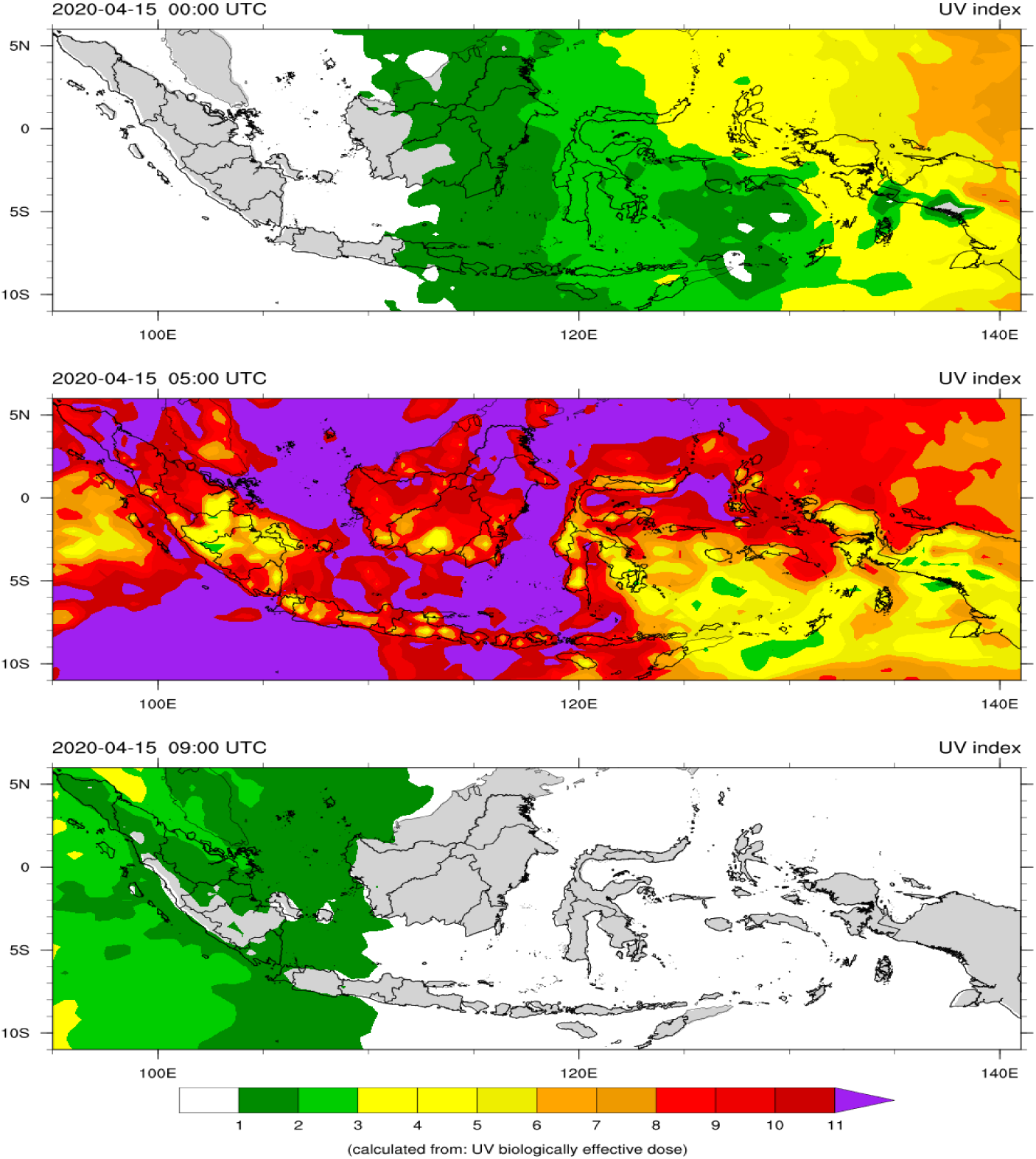
The spatial variation of UV index calculated from the ECMWF’s uvbed product for April 15, 2020.

### 1.3. UV index model vs observation: a comparative analysis

In this study, the two ECMWF UV products were used as a comparison to the ground-based data collected from the three locations for the period of January to March 2020. After calculating the UV index from both observation and model data sets, daytime hourly UV indices were analyzed to indicate whether there is an agreement between the two data sets. The comparison from the three locations can be seen in fig. 2b - d.

**Figure 2:**
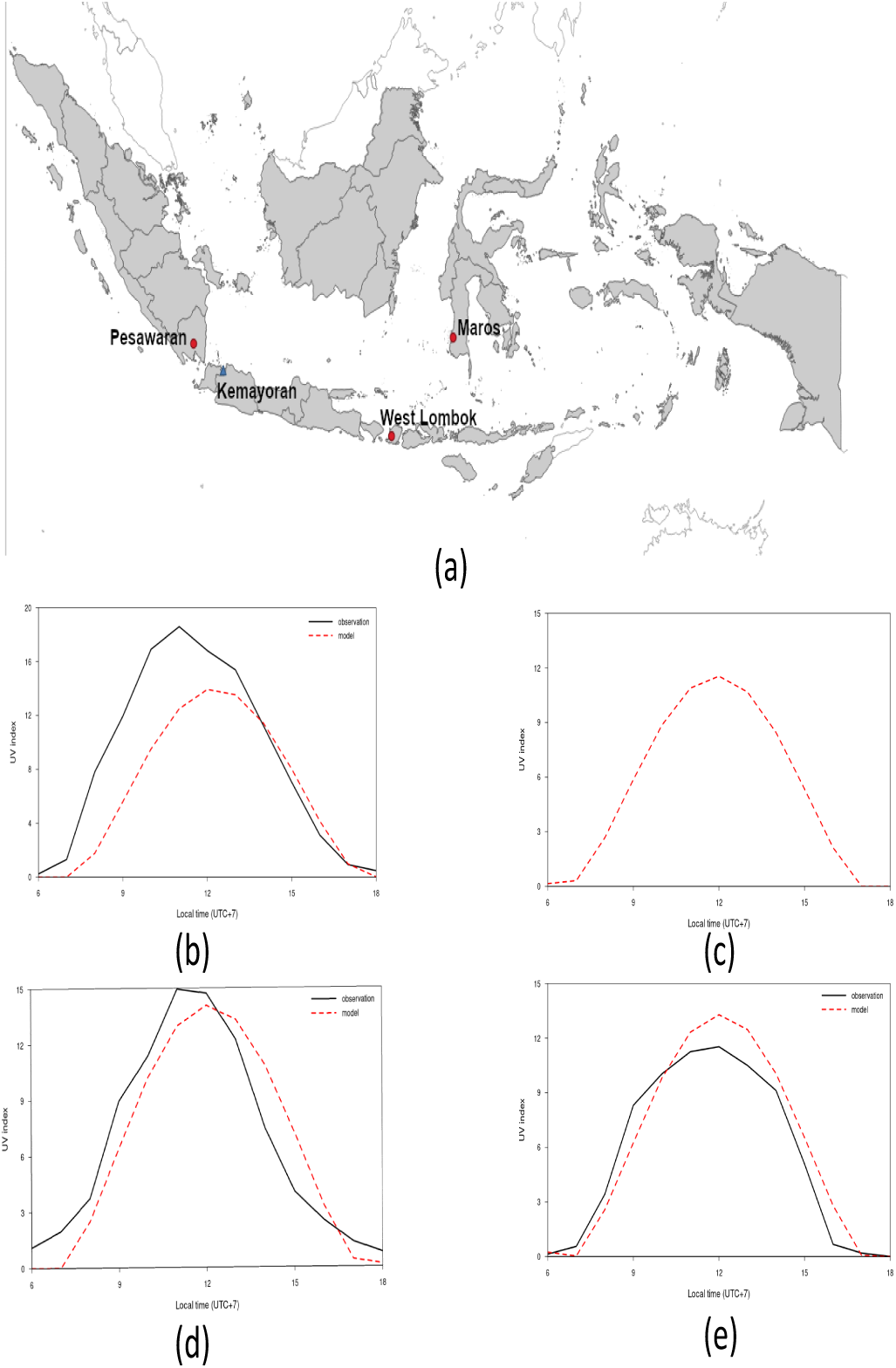
(a).ground-based data collected from the three locations (Pesawaran, Maros, and West Lombok) for the period of January to March 2020 (b). mean hourly UV indices in West Lombok Province. (c). T mean hourly UV indices in Kemayoran, Jakarta Province. (d). mean hourly UV indices in Maros, South Sulawesi. (e). mean hourly UV indices in Pesawaran, Lampung Province.

Fig. 2e and 2d show mean hourly UV indices in Pesawaran, Lampung Province, and in Maros, South Sulawesi Province, respectively. It can be observed that, in general, the model is able to capture the hourly pattern of UV indices calculated from measured UV radiation. There are some slight differences on the estimated UV indices, which might be attributed to the coarse horizontal resolution of the model that did not include the dynamics of atmospheric conditions on a finer scale. Nevertheless, these results can be considered as a good agreement between the observation and the model.

A different result, however, is shown in 2b, where West Lombok in West Nusa Tenggara Province observed the UV index difference between the model and the observation. During its peak, the model underestimates the observation by more than five units. There are two factors that might be contributed to this difference. Firstly, the model was not able to capture the local characteristics of UV radiation received on the surface in this location. These local characteristics can be associated with the topographical features and the predominant weather conditions in this location. Secondly, the selection of weighting factors to adjust UV-A and UV-B radiation is not suitable for this location. It is possible that the UV radiation received on the surface is at different wavelengths from one place to another. The absorption of UV radiation is governed by atmospheric compositions and weather conditions. Therefore, the amount of energy brought by UV radiation is subject to at what wavelength the radiation is able to reach the surface. This result highlights the importance of future studies that involve more precise measurements on UV radiation from different wavelengths. These studies are required to determine the prevalent wavelength of UV radiation that is measured on the ground.

Notwithstanding the disagreement between the model and the observation shown in 2b, these results suggest an encouraging finding that the model may be suitable for estimating the UV index in locations where there is no ground-based data.This is of particular importance because of the limitation of measurement availability in Indonesia. However, this method and its interpretation have to be approached with cautions because of there is still a need to get more information on the prevalent UV wavelength discussed earlier.

Next, we look into the pattern of daily confirmed cases and UV index in several countries. As a remainder, UV index can be measured as below:

- 0 to 2: Low Risk of harm from unprotected sun exposure.
- 3 to 5: Moderate Risk of harm from unprotected sun exposure.
- 3 to 5: Moderate Risk of harm from unprotected sun exposure
- 6 to 7: High Risk of harm from unprotected sun exposure
- 8 to 10: Very High Risk of harm from unprotected sun exposure
- 11+: Extreme Risk of harm from unprotected sun exposure

It possibly harms to people that are not used to live in such an extreme UV index [17], however, it can reduce virus activity. The people who live in high UV index areas such as Africa and other tropical countries have already long-adapted in such situation. They may take advantage that pandemics are not as excessive as subtropical countries.

### 1.4. Difference UV between tropical location and covid-19 pandemic growth

Fig. 3a shows uv index over the Asia and Europe on March 28 2020 that seems to be correlated to covid-19 pandemic rate over countries on April 4 2020 (3b). Most of high rate of covid-exposed countries are located in subtropical area, conversely, low rate of covid-exposed countries are spreading throughout tropical areas. Concurrently, tropical countries have high index of UV index (> 7) over time whilst subtropical countries are interchanging between low (< 4) and high index (> 7) of UV index depending on the season. The confirmed cases map in Fig.la seems to be correlated to UV index map where the majority of confirmed people live in the northern subtropical countries (fig. 3b). The southern subtropical countries are still in low rate of pandemic growth, however, the earth is revolutionize and between May to August they are gradually entering cold season with low level pf UV index. They have advantages of early anticipation of covid-19 growth in situation where UV index is still high.

**Figure 3:**
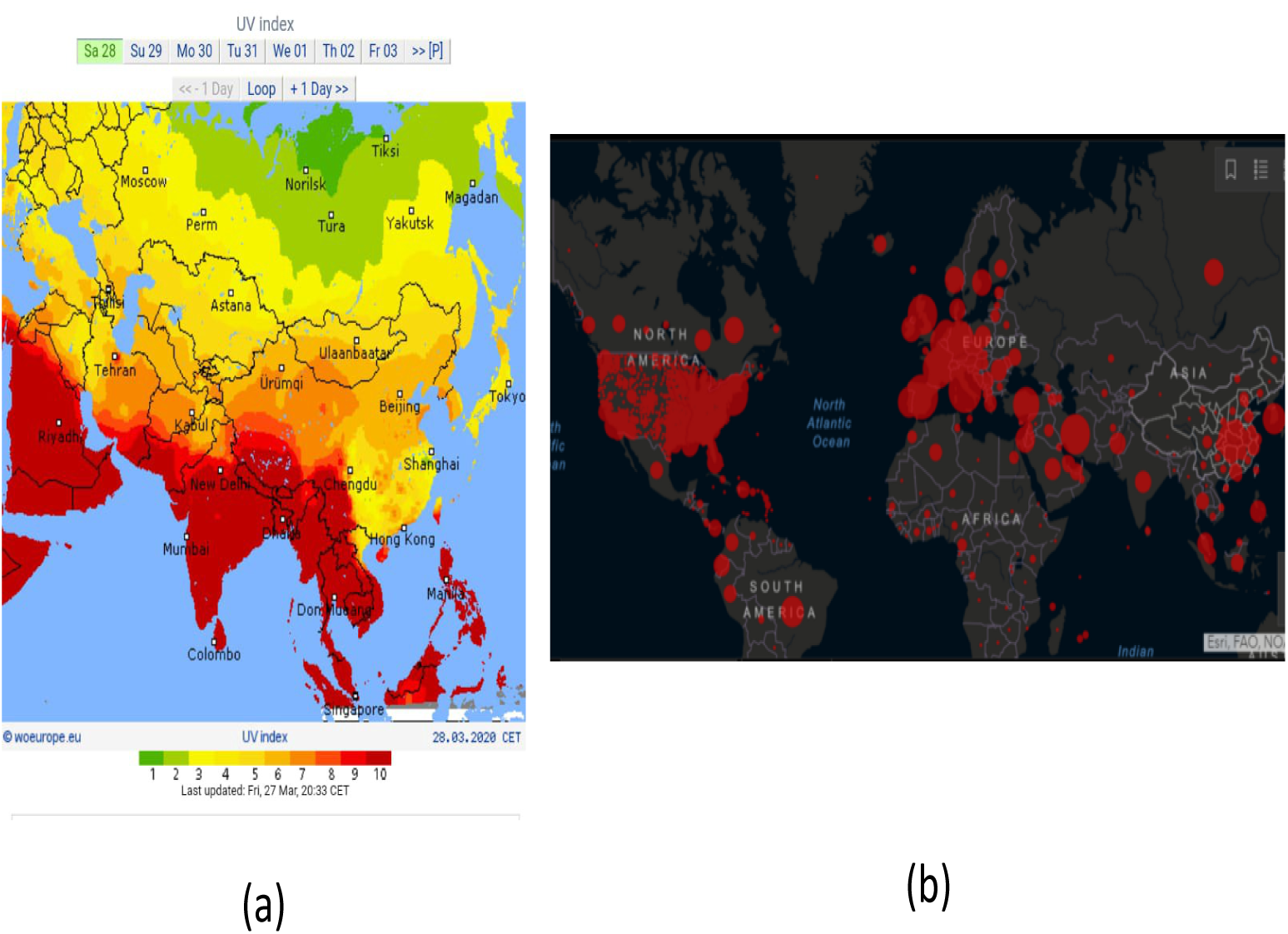
(a).Visualization of uv index distribution over continents (woeurope.eu). (b). Visualization of conformed covid-19 cases over continents (John Hopkins University Medicine on April 4, 2020).

### 1.5. How coronavirus spreads differently in each country

Coronavirus which causes the illness known as covid-19 was first reported in China in December 2019. By March 28 2020, this disease has been spreading to at least 178 countries and territories. In some countries, the accumulation of confirmed cases varies from 0 to more than 120000 people over time (fig. 4a and 4b). This data shows that this virus has the ability to spread easily and quickly. Indonesia as one of the countries affected by coronavirus shows the number of cases climbing to 1000 people. The number seems to be high but it is still lower when compared to other countries such as America, Italy, France, Netherlands, and Iran.

**Figure 4:**
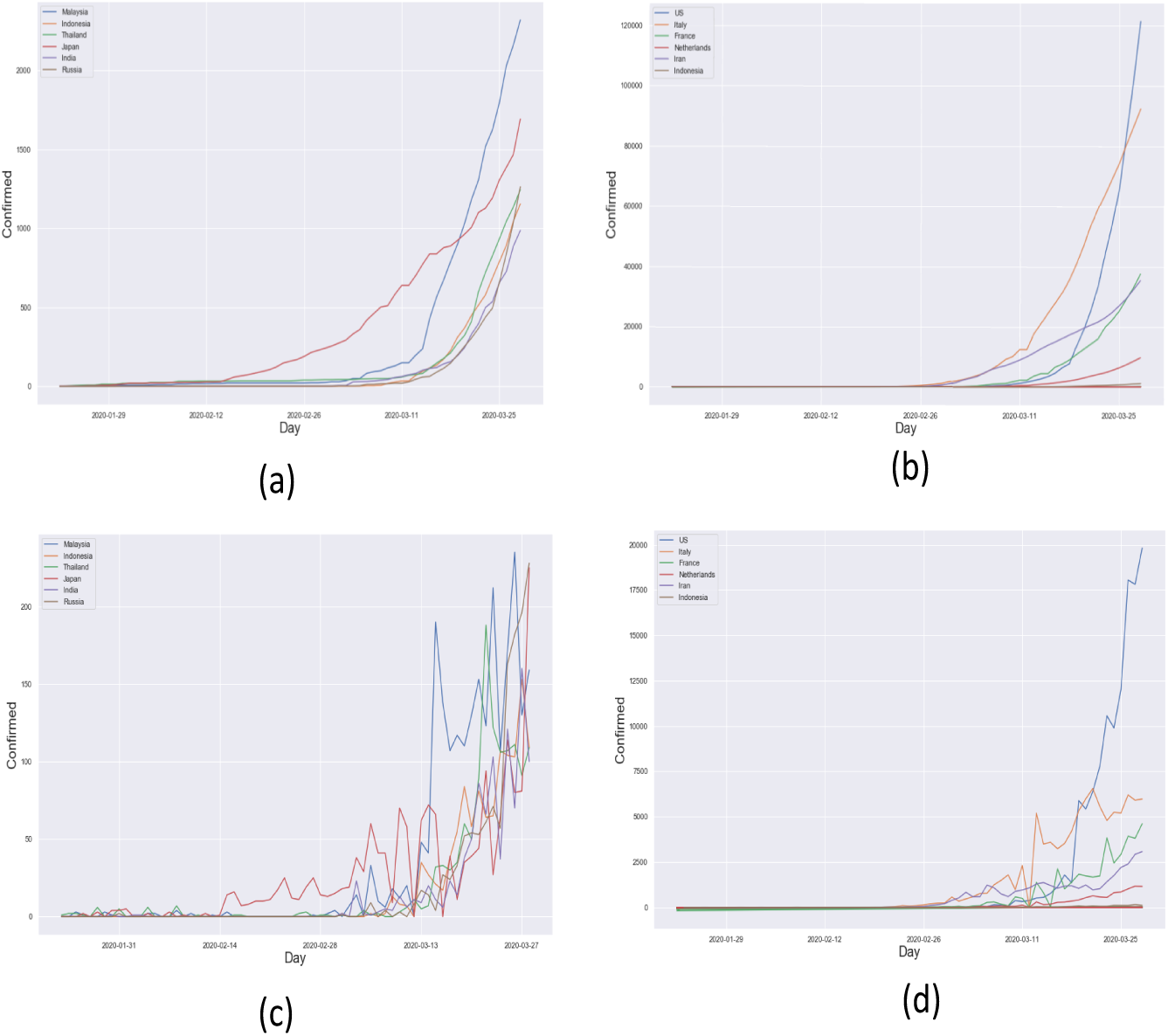
(a). The pandemic growth of countries which have low level of growth rate. It is shown that confirmed accumulation in range of 0–2000+ people. (b). The pandemic growth of countries which have low level of growth rate. It is shown that confirmed accumulation in range of 0–120000+ people. (c). Daily confirmed case in low growth rate countries. (d). Confirmed case per day in some countries

Fig. 4a and 4b show the spread of coronavirus in some countries that less than 5000 confirmed cases and more than 5000 confirmed cases, respectively. The accumulation of confirmed case grows exponentially with different rate in each country. Indonesia grows in very little rate compared to US, Italy, France, Netherlands, and Iran. We can easily cluster those countries by its location using pandemic growth information. For example, referring to Fig. 3a and 3b, Malaysia, Indonesia, Thailand, and India can be grouped into one cluster while US, Italy, France, Netherlands, Iran, Japan, and Russia in another cluster by its tropical and subtropical continent, respectively. Japan, however, has higher UV index compared to US, Italy, and Russia and its UV index grows over time as spring season is coming (fig. 5a). Russia that has low population density seems to have anomaly here and of course another parameter such as humidity, air pollution or even economic relation with epicentre country can be taken into account in future studies (update: by May 22 2020, the total of conformed cases in Russia is positioned at second after US).

**Figure 5:**
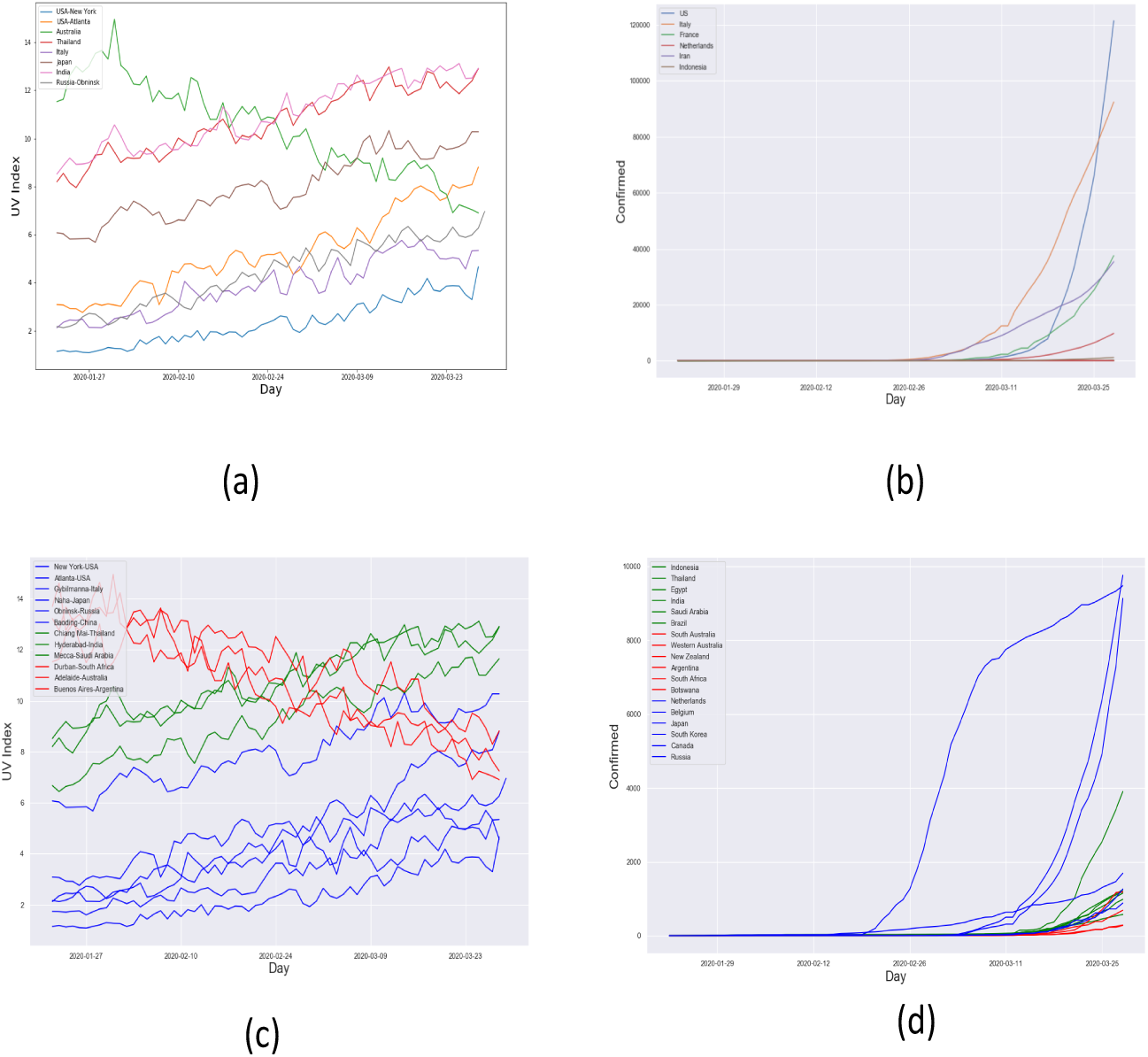
(a).UV Index over time in many countries. (b). The pandemic growth of countries. (c). The UV index alteration over time in northern sub tropical countries (blue), tropical countries (green), and southern sub tropical countries (red). (d). The covid-19 pandemic growth in northern sub tropical countries (blue), tropical countries (green), and southern sub tropical countries (red). Note that US, UK, Italy, or Spain is not included due to very high growth rate.

### 1.6. Daily cases vs UV index

Before, let us show the covid-19 epidemic growth in several countries and how exponential its slope between January 22 2020 to March 28 2020. The number of confirmed cases of people infected with the corona virus tends to increase every day (fig. 4c and 4d). This trend shows how quickly this virus transmits among humans. Inside human body, this virus takes time to show symptoms of illness. it makes coronavirus easily spreading among people before the carrier notice about it. Highest confirmed case reported in US reaches around 20000 people in a day on March 28 2020 (Fig. 4d).

Based on the fig. 5a, during the same days with pandemic growth in fig. 5b, northern subtropical countries have UV index grows over time as the cold season ended and spring season is coming. Australia as a representation of subtropical countries has its UV index decreasing over time as summer season ended and fall season is coming. In Atlanta, area which is located in central area of US, has higher UV index than of in New York, even though they both grow concurrently over time. Interestingly, New York area has lowest UV index compared to others of which has UV index of 1 to 4 over time. In tropical countries, such as Thailand and India, UV index tends to be stable over time during January 22 2020 to March 28 2020.

### 1.7. Tropical vs Subtropical: interesting characteristics

Fig.5d shows pandemic growth in tropical (green), northern subtropical (blue), and southern subtropical (red). It is quite interesting that the blue countries grow exponentially over time since the initially confirmed people are recorded, with sharper, and faster than the green and red countries. The green countries grow sharper and faster than red countries, even though the growth of some countries has cross points with each other. The overtaking points indicate that there exist growth pace that is becoming slower than the other, and vice versa. This happens between two adjacent groups either blue with green or green with red. This phenomenon possibly can be explained in Fig.5c.

Fig.5c shows the change of UV index over time in northern subtropical countries (blue), tropical countries (green), and southern tropical countries (red). We can easily understand that the blue and green countries are mono-tonically increasing over time while the red countries are monotonically decreasing. This phenomenon regards the changing season between cold to summer in northern countries and, conversely from summer to cold season in southern tropical countries. The green countries rarely have UV Index below 6. UV Index behavior of blue, green, and red countries are concurrent with the accumulation of confirmed covid-19 over time in Fig.5d. Some blue countries are starting to be sloppier as higher UV Index and with proper social distancing.

Even though we still do not know the final growth curve, by the aforementioned evidence, they should prepare the worst case. The dynamic patterns of the current situation show that some southern subtropical countries are starting to grow exponentially as shown in Fig.5d. Especially, for airborne covid-19 viruses that exist in the air, spreading vastly in the air. They can perform early restrictions before exponential increase whilst also reduce pollution to let UV lights reach the surface.

### 1.8. China case: recovering with the help of UV light and lockdown?

Fig.6a shows graphs of confirmed case accumulation over time in China. There is an interesting fact that Hubei province in which Wuhan city is located does not extremely spread covid-19 to other provinces. Fig. 6b shows graph of daily confirmed cases in China. There is also an interesting fact that Hubei province, in which Wuhan city is located, does not extremely spread covid-19 to other provinces. The daily confirmed cases show that other provinces do not show significant confirmed cases compared to Hubei province. This is related to China government effort to carry out tight lockdown in the epicentre (Hubei area) thorough screening, testing and contact tracing programs, as well as bringing in early social distancing whilst also light lockdown in another area [18]. The reason can be two-fold: the help of lockdown with its social distancing and by the help of increasing UV index as shown in Fig.6c. All four stations of Tianjin, Dalian, Mount Waliguan, and Baoding show increased UV index over time during January 22 2020 to March 28 2020.

**Figure 6:**
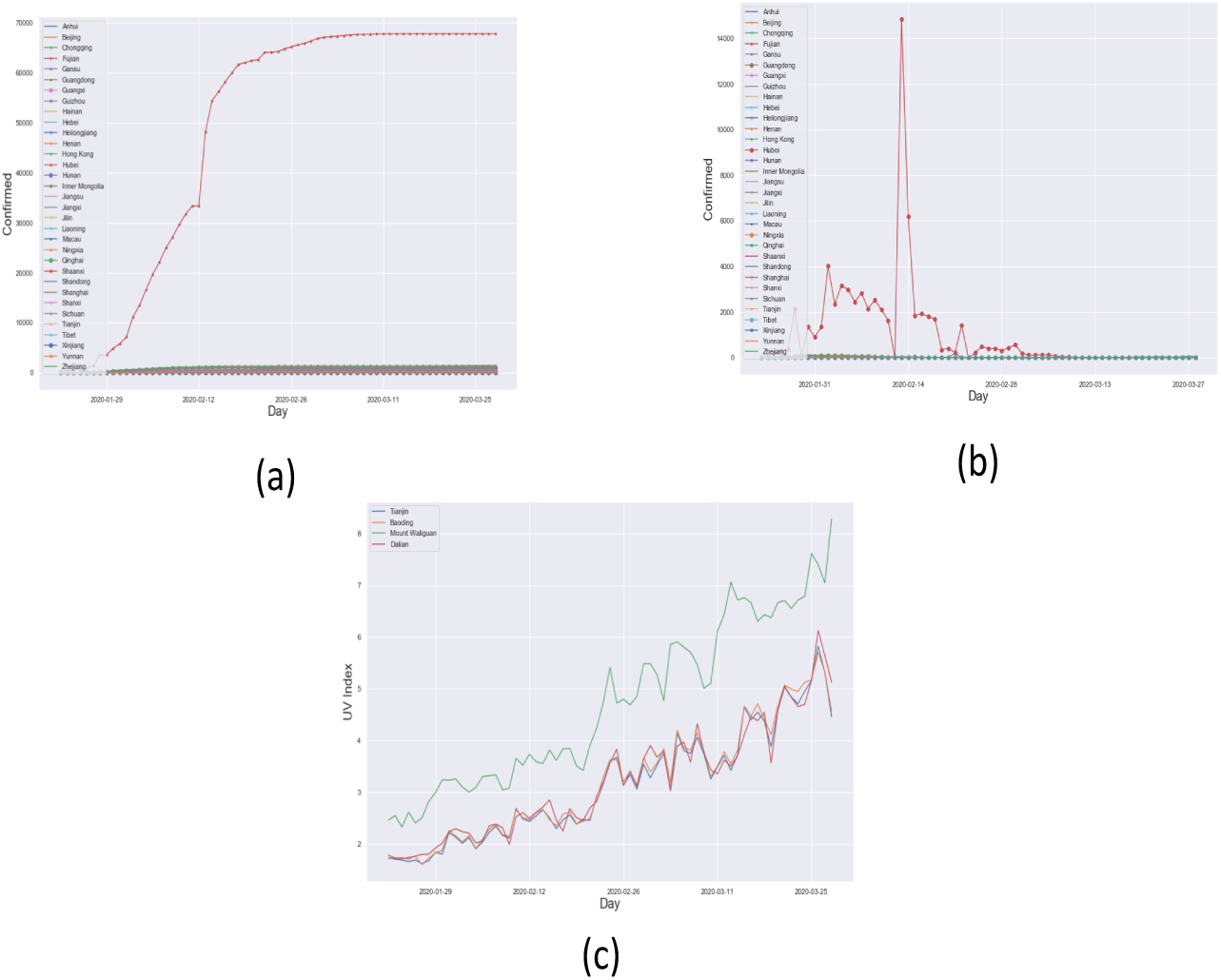
(a) Accumulation of confirmed case in China until the days of growth halt. (b). Daily confirmed case in China. (c). The UV index over time in China. The four stations are presented which are Tianjin and Dalian in northern China, Mount Waliguan in central China, and Baoding represents middle and southern China.

### 1.9. Growth Slope: see the change over time

We use local and global slopes by means tangent line [7] as parameters to see how related the UV index time series with the accumulation of confirmed covid-19 growth as shown in fig. 7. tangent line is used as the steepness metric of the graph. The tangent line is defined as follows:

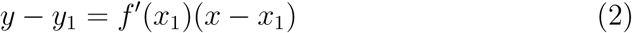

**Figure 7:**
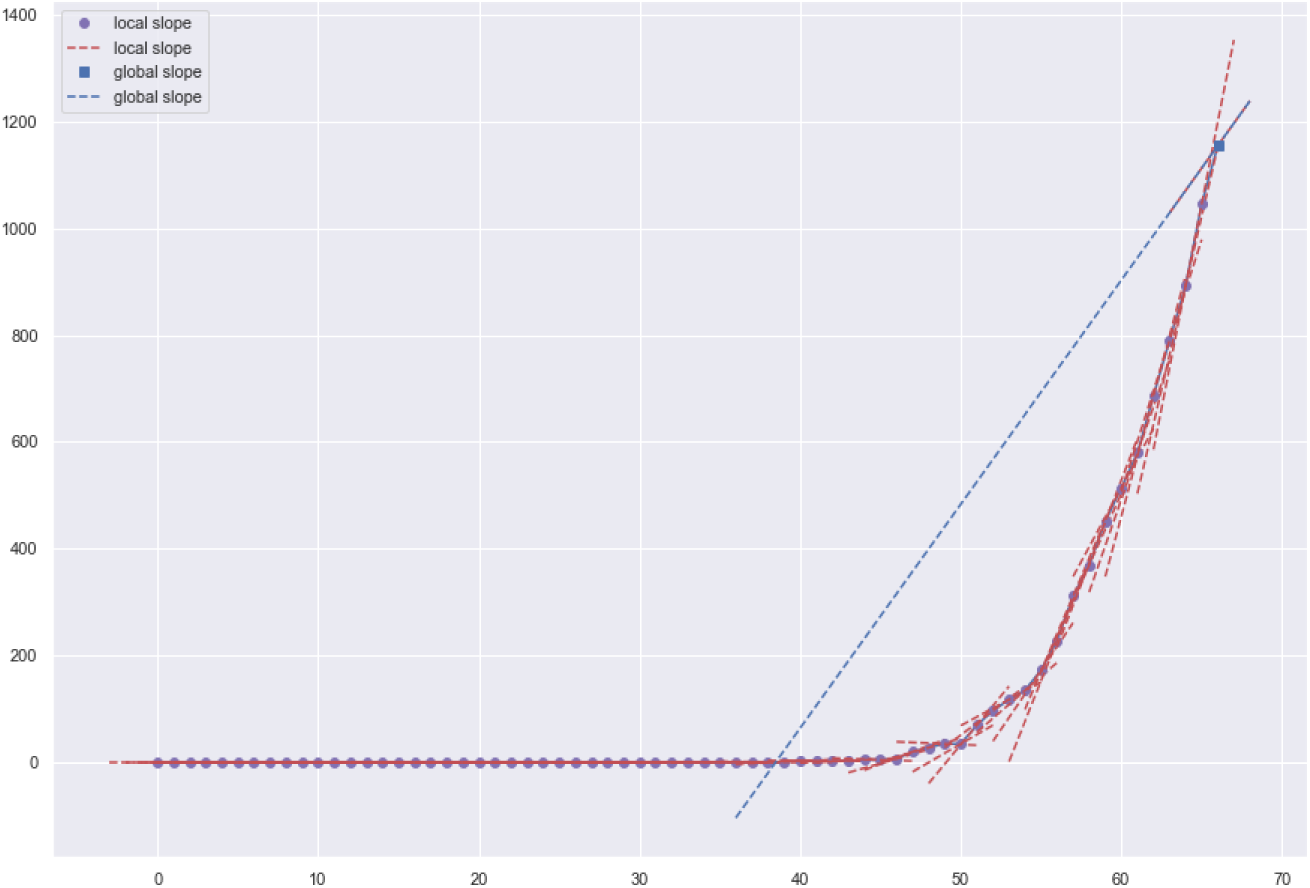
Example of local slopes and global slope

Where *f* is growth function and *x* and *x*_1_ are target and initial point, respectively. The difference between local and global slope is that local slope uses each local point in time frame relatives to initial point *y*_1_ to estimate the tangent line while the global slope is measured from the last point relatives to initial point *y*_1_. The global slope gives more global growth performance.

### 1.10. Correlation Test: to see the relationship between parameters

Let me remind you that:

- 1.0 = positively correlated; for instance, if A increases, B also increases, and vice versa.
- 0.0 = no correlation; for instance, if A increases, B does not change, and vice versa.
- -1.0 = negatively correlated; for instance, if A increases, B also decreases, and vice versa.

Let us introduce you to several parameters to be tested:

- UVIEF: cloud-free,erythemal (sunburn),UV index.
- UVDEF: cloud-free,erythemal (sunburn),UV dose,[kJ/m2].
- UVDDF: cloud-free,dna-damage,UV dose,[kJ/m2].
- UVDEC: cloud-modified,erythemal (sunburn),UV dose,[kJ/m2].
- ozone: local,solar,noon,ozone,column,[DU].

UV index is a measure for the effective UV irradiance (1 unit equals 25 mW/m2) reaching the Earth’s surface in clear sky. UV dose is the effective UV irradiance (given in kJ/m2) reaching the Earth’s surface integrated over the day and taking the attenuation of the UV radiation due to clouds into account. Total column ozone is the total amount of ozone in a column extending vertically from the earth’s surface to the top of the atmosphere. It is measured using ground-based stations and satellites and is reported in Dobson units (DU). The ozone hole is defined in terms of reduced total column ozone — less than 220 DU.

The correlation map between variables is defined as follows:

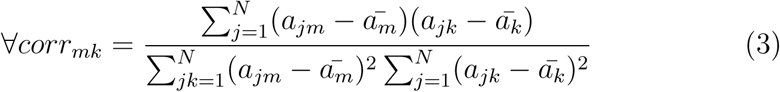

Where for all available variables we calculate correlation between variables *m* and *k*.

### 1.11. Correlation test of the world data

Countries to be included are Australia, Thailand, India, Japan, US, Italy. Global Atmosphere Watch (GAW) station to be used:

- AcadiaNatForest, USA
- Gibilmanna, Italy
- Chiang Mai, Thailand
- Hyderabad, India
- Adelaide, Australia
- Naha, Japan

Fig.8a shows correlation map between parameters of the world. Average of UV index and average of ozone over time are correlated to global slope of confirmed cases accumulation by −0.86 and 0.94, respectively. Meaning that the higher UV Index is, the lower the growth rate of covid-19 pandemic would be, and vice versa with a correlation of −0.86. Conversely, the higher the ozone value is the higher growth rate of covid-19 pandemic would be, and vice versa with a correlation of 0.94 as shown in fig. 8a. the global slope of UV index alteration over time is correlated to the global slope of confirmed cases accumulation by 0.7. This is understandable since the UV index in northern subtropical countries tends to increase over time indicated by high tangent value, however, the growth rate of covid-19 pandemic is still increasing even though the pace is slowing down. Conversely, in southern subtropical countries, the UV index tends to decrease over time making the tangent value to be low even minus. We should wait for the next several months to see whether southern subtropical countries will exponentially increase the confirmed cases or not as data taken only until March 28, 2020.

**Figure 8:**
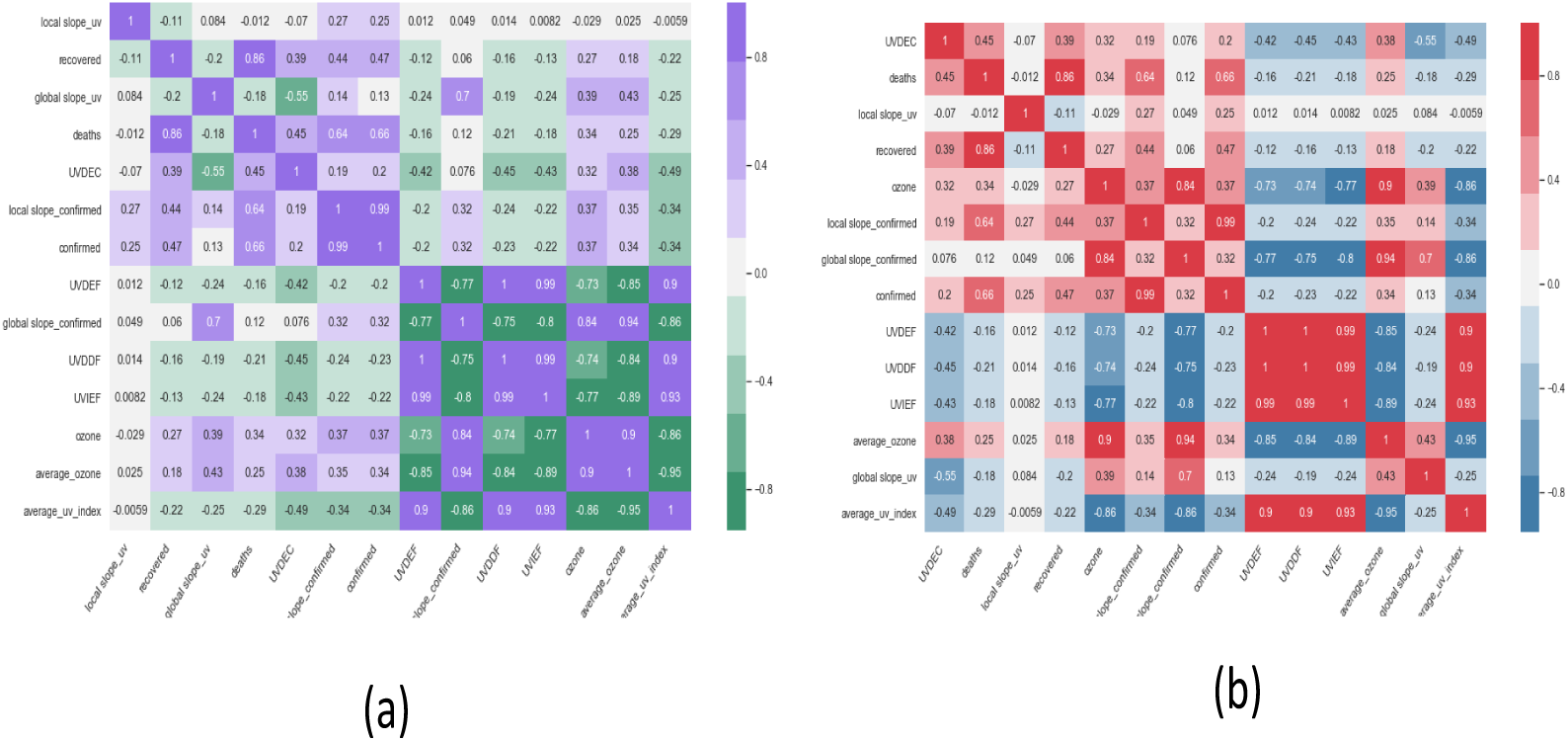
(a) Correlation map between parameters of the world. (b). Correlation map between parameters of China.

### 1.12. Correlation test of China data

Provinces to be included are Tianjin, Hebei, Liaoning, Qinghai. GAW station to be used:

- Tianjin
- Baoding
- Mount Waliguan
- Dalian

Fig. 8b shows correlation map between parameters of China. Average of UV index and average of Ozone over time are correlated to global slope of confirmed cases accumulation by −0.86 and 0.94, respectively. Meaning that the higher UV Index is, the lower growth rate of covid-19 pandemic would be, and vice versa with a correlation of −0.86. Conversely, the higher the ozone value is the higher growth rate of covid-19 pandemic would be, and vice versa with a correlation of 0.94 as shown in fig. 8b.

## 2. Discussion

Based on these findings, anticipation in form of gradual from loose to tight in southern subtropical countries in accordance with UV index over time might be taken into account to reduce the economic burden. Tropical countries might take advantage of its high UV Index in the entire session but has to keep anticipating. Special attention in the central economy area where there is heavy air pollution should be considered because the high UV index does not have enough potential to inactivate the virus. Meaning that it should be anticipated that the exponential phase will be high.

As we can see in fig. 9, Jakarta, Bangkok, Wuhan, Beijing, and Milan have higher pollution of PM2.5 than Singapore, Tokyo, Brussels. However, Sydney and Adelaide have the lowest PM2.5 pollution. As remainder, PM2.5 is kind of particulate matter produced from diesel exhausts, home cooking, smoke from burning wood, etc. It is measured by microgram per cubic meter (Fig. 6). Low pollution corresponds to the best air quality index within the range of January-April and concurrently with pm2.5 pollution rate, Australia has the lowest rate of covid-19 exposure. Now, we try to correspond with data of UV light time series within January to April. Sydney and Adelaide have a high UV index while their pollution is at the lowest level compared to others of which concurrent with low confirmed covid-19 cases in Australia. Moreover, they also did an early anticipation by holding initial social distancing and another restrictions to facilitate medical experts to test, find, quarantine, and handle suspects as soon as possible. In northern subtropical cities, Brussels has low both UV index and pollution. In other hand, Milan (Italy) has high pollution whilst at the same time its condition is worsened with low UV index during that season, January until April. Tokyo has a moderately low UV index and pollution. In tropical city, Singapore has a moderately high UV index and pollution, however, remember that they have done tests to 20815 / 1 million population dated on April 12 2020, a higher sampling rate than the others. In China, from January to February, Wuhan and Beijing have a high pollution level and gradually become lower starting from March to April, therefore, they have a longer standard deviation of average of pm2.5 than the others. China is also recorded to have an upward UV index within the range of February-April based on UV light time series data in Fig. 6c. Another tropical cities, Jakarta and Bangkok have a high pollution rate with mean of pm2.5 rate is around 100, similar to Wuhan and Beijing, however, the UV index of Indonesia at that time is slightly lower than Thailand which regards the confirmed covid-19 case of Thailand is less than Indonesia even though located in the same tropical countries. Note that the epicenter of Indonesia and Thailand is Jakarta and Bangkok, respectively.

**Figure 9:**
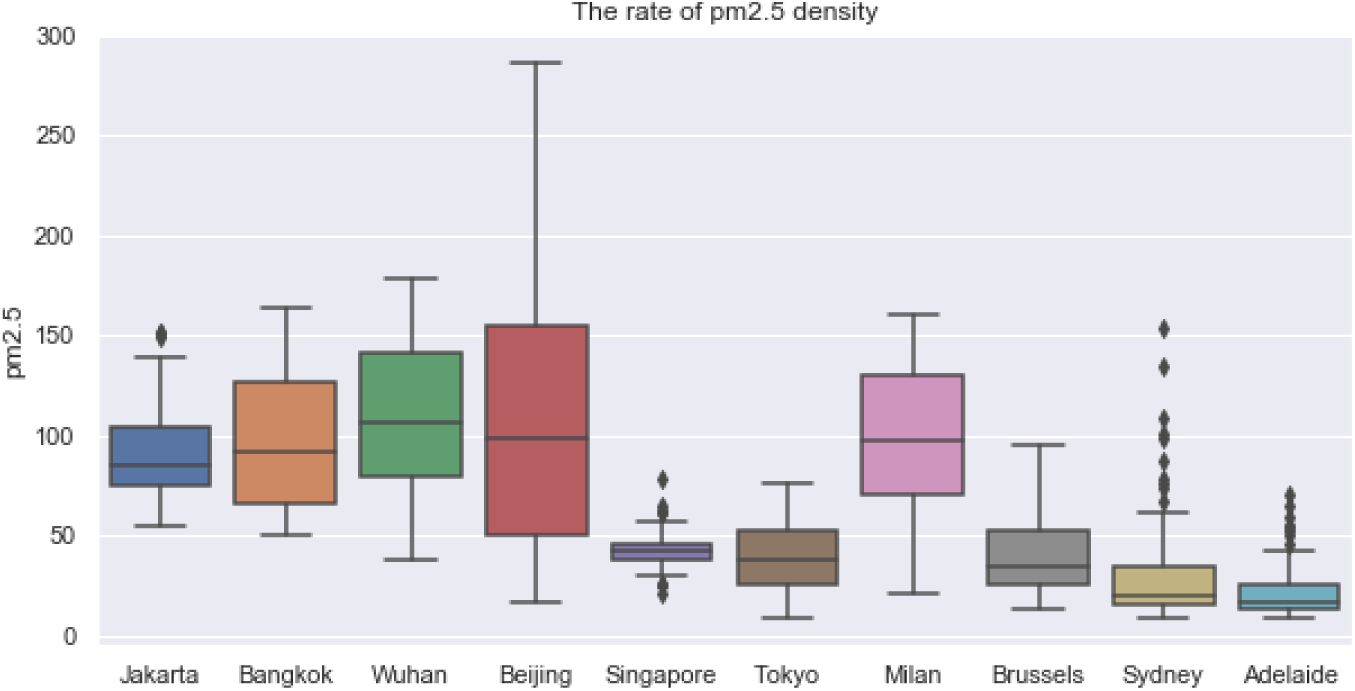
pm2.5 boxplots of epicenter cities during January to April.

Now, we look into regression analysis between PM2.5 rate and total confirmed covid-19 cases on April 12 2020. We split cities to tropical and southern subtropical cities (fig. 10a) and northern subtropical cities (fig. 10b). This split follows assumption of UV index cluster during January to April. The regression of tropical cities in fig. 10a shows that they are positively correlated. It is shown that we have an outlier of Jakarta, Singapore, Hanoi, and Ho Chi Minh, however, Jakarta and Singapore are still higher than of Adelaide, Sydney, and Auckland of which located in southern subtropical countries. The problem of a developed country that confidently located in the northern subtropical area is the high level of pm10 of which usually product of factories. However, in cities, the problem of pm2.5 might still exist. fig. 10b shows that Milan, Wuhan, Madrid, Los Angeles are in the same cluster though it still has outliers of Seoul and Madrid. Chicago, Los Angeles, Detroit, Berlin, Tokyo, and Amsterdam in another cluster. This result is in line with number of confirmed covid-19 case where Wuhan, Milan, and Los Angeles have higher accumulation number of confirmed covid-19.

**Figure 10:**
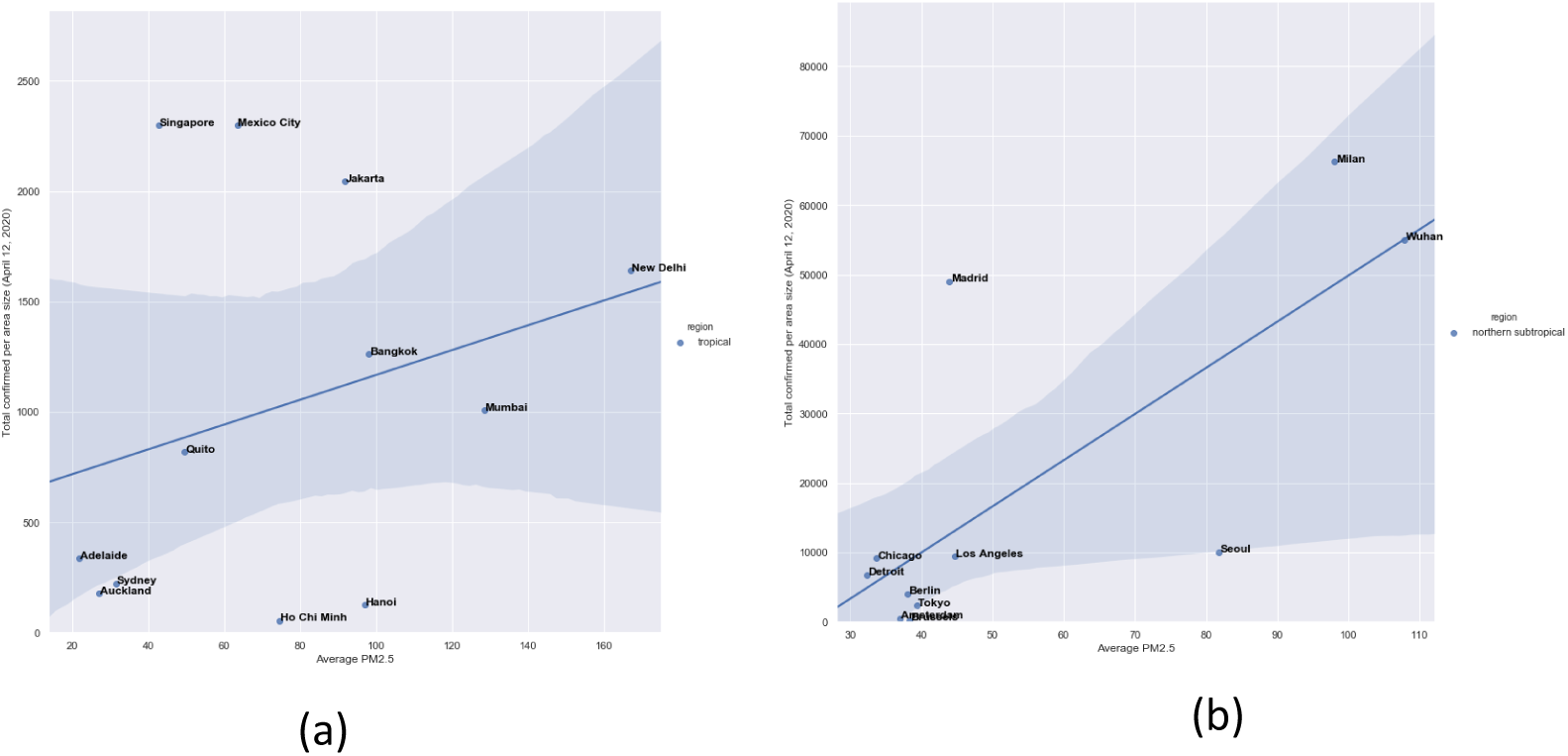
Regression analysis between confirmed cases and average of PM2.5 in (a) tropical and southern subtropical cities and (b) northern subtropical cities.

Temperature and humidity possibly influence the covid-19 spread [19]. It was found that greater survival of coronavirus occurs in lower temperature and humidity. Based on its results, it is indicated that geographical locations where higher temperature and humidity exist such as tropical countries have advantages in withstanding the covid-19 spread. Humidity has greater role in inactivating virus than temperature either on surface or in the air. It could explain why some tropical countries has more confirmed covid-19 cases than the others. These conclusions open up future research potential where UV, pollution, and humidity can be strong features to predict and analyse the spread of covid-19.

## 3. Conclusion

While UV index and ozone are correlated with global spread of corona pandemic, we believe it is not just a standalone factor. There are some other factors such as economic activity and population density influencing the spread and growth of coronavirus cases. However, UV light and ozone are strong enough to be taken into account to minimize the global covid-19 pandemic effect. In the cities where there is heavy air pollution, the high UV index might be no meaning in terms of inactivating the virus. Next, we would like to investigate on a smaller scale in one nation such as Indonesia with humidity and pollution are included. Another thing is we are preparing to predict when pandemics will end in each country using neural network model.

## Data Availability

Data are publicly available and referred in manuscript

https://www.github.com/datasets/covid-19/tree/master/data

https://www.temis.nl/uvradiation/UVarchive/stations_uv.html

https://www.aqicn.org

https://github.com/cbasemaster/uvcorona

## References

[1] C. Wang, P. W. Horby, F. G. Hayden, G. F. Gao, A novel coronavirus outbreak of global health concern, The Lancet 395 (2020) 470–473.

[2] D. Hui, I azhar e, madani ta, et al. the continuing 2019-ncov epidemic threat of novel coronaviruses to global health-the latest 2019 novel coronavirus outbreak in wuhan, china, Int J Infect Dis 91 (2020) 264–266.

[3] N. Singh, M. Kaur, On the airborne aspect of covid-19 coronovirus, arXiv preprint arXiv:2004.10082 (2020).

[4] N. Madhav, B. Oppenheim, M. Gallivan, P. Mulembakani, E. Rubin, N. Wolfe, Pandemics: risks, impacts, and mitigation, in: Disease Control Priorities: Improving Health and Reducing Poverty. 3rd edition, The International Bank for Reconstruction and Development/The World Bank, 2017.

[5] E. Callaway, D. Cyranoski, S. Mallapaty, E. Stoye, J. Tollefson, The coronavirus pandemic in five powerful charts., Nature 579 (2020) 482–483.

[6] M. M. Jensen, Inactivation of airborne viruses by ultraviolet irradiation, Appl. Environ. Microbiol. 12 (1964) 418–420.

[7] A. N. Strahler, Quantitative slope analysis, Geological Society of America Bulletin 67 (1956) 571–596.

[8] K. Bergmann, Uv-c irradiation: A new viral inactivation method for biopharmaceuticals. american pharmaceutical review. consultado el 01/04/2020, 2014.

[9] P. Koronakis, G. Sfantos, A. Paliatsos, J. Kaldellis, J. Garofalakis, I. Koronaki, Interrelations of uv-global/global/diffuse solar irradiance components and uv-global attenuation on air pollution episode days in athens, greece, Atmospheric Environment 36 (2002) 3173–3181.

[10] W. F. Barnard, B. N. Wenny, Ultraviolet radiation and its interaction with air pollution, in: UV Radiation in Global Climate Change, Springer, 2010, pp. 291–330.

[11] R. Kuznia, The timetable for a coronavirus vaccine is 18 months. experts say that’s risky, 2020. URL: https://edition.cnn.com/2020/03/31/us/coronavirus-vaccine-timetable-concerns-e

[12] T. Koutchma, Advances in ultraviolet light technology for non-thermal processing of liquid foods, Food and Bioprocess Technology 2 (2009) 138–155.

[13] K. N. Prodouz, J. C. Fratantoni, E. J. Boone, R. F. Bonner, Use of laser-uv for inactivation of virus in blood products (1987).

[14] J. W. Tang, The effect of environmental parameters on the survival of airborne infectious agents, Journal of the Royal Society Interface 6 (2009) S737-S746.

[15] J. Van Geffen, R. Van Der A, M. Van Weele, M. Allaart, H. Eskes, Surface uv radiation monitoring based on gome and sciamachy, 2004.

[16] A. McKinlay, B. Diffey, W. Passchier, Human exposure to ultraviolet radiation: risks and regulations, Excerpta Medica, Amsterdam, Netherlands (1987).

[17] K. Ezzedine, C. Guinot, E. Mauger, T. Pistone, M.-C. Receveur, P. Galan, S. Hercberg, D. Malvy, Travellers to high uv-index countries: Sun-exposure behaviour in 7822 french adults, Travel medicine and infectious disease 5 (2007) 176–182.

[18] Z. Wu, J. M. McGoogan, Characteristics of and important lessons from the coronavirus disease 2019 (covid-19) outbreak in china: summary of a report of 72 314 cases from the chinese center for disease control and prevention, Jama 323 (2020) 1239–1242.

[19] L. M. Casanova, S. Jeon, W. A. Rutala, D. J. Weber, M. D. Sobsey, Effects of air temperature and relative humidity on coronavirus survival on surfaces, Appl. Environ. Microbiol. 76 (2010) 2712–2717.

